# Safely Reopening K-12 Schools During the COVID-19 Pandemic

**DOI:** 10.1101/2020.10.07.20208710

**Authors:** Courtney D. Shelley, Philippa S. Chadwick, Carrie Manore, Sara Y. Del Valle

## Abstract

Early school closures were a consistent, nationwide response to the COVID-19 pandemic in mid-March due to the role that children play in spreading influenza. This left us with limited understanding of COVID-19 transmission in children until several states reopened schools for the 2020-2021 school year. While early school closures were likely beneficial in protecting children in the initial stages of the pandemic in the U.S., long-term closures pose significant cumulative effects in children who rely on schools for instruction and additional social services, and for parents who need to balance work and childcare obligations. Reopening schools safely is a high priority for many interested stakeholders.

Proposed in-person school reopening plans include traditional, 100% school capacity, five days per week instruction, hybrid scenarios with reduced in-person instruction and virtual learning, and various reduced school capacity schedules with non-pharmaceutical interventions in place. To assess the potential impacts of different reopening plans, we created a modified SIR-type transmission model that captures multiple known pathways of COVID-19 transmission in a 100,000-person community.

Our results show that plans that utilize consecutive days in school and divide students into separated smaller cohorts who attend school together, as well as plans that emphasize distance learning, are better able to suppress disease spread and reduce risk from an introduced infective into the community. Plans with more consecutive school days are protective for both the schoolchildren and surrounding community by acting to separate the larger intermixing population into smaller intermixing subpopulations. The “Five-Day Switch” plan, which separates students into two cohorts, each of whom attend in-person learning for five consecutive days followed by five days of distance learning, best captures these protective attributes. All modeled plans assumed initially disease-free communities and that children’s interactions with the community are greatly reduced during instructional days, both for in-person and distance learning.

## INTRODUCTION

A cluster of idiopathic pneumonia cases in Wuhan, Hubei Province, China was reported via the Program for Monitoring Emerging Diseases (ProMED) on 30 December 2019 (ProMED 2019). The viral agent was identified by Chinese health authorities as a novel coronavirus on 9 January 2020 (World Health Organization 2020a) and officially named SARS-CoV-2 (World Health Organization 2020b). By 20 January 2020, there were 9,976 cases of coronavirus disease (COVID-19), including the first reported U.S. case (Holshue et al. 2020). The World Health Organization issued a statement on 7 March 2020 in recognition of 100,000 COVID-19 cases worldwide. Within a week, many U.S. schools closed for an early Spring Break, with most schools remaining closed for the duration of the 2019-2020 school year. This measure was in part based on knowledge of the spread of influenza, where children play a key role in disease transmission (Isfeld-Kiely and Moghadas 2014, Bayham and Fenichel 2020, Russell et al. 2016).

Data on children’s susceptibility to COVID-19 has been inconsistent, especially due to early school closures as one of the only consistent and prompt national actions in response to the U.S. pandemic (Auger et al. 2020). Early international evidence showed children were significantly less likely than adults to become infected (Davies et al. 2020, Gudbjartsson et al. 2020). The Centers for Disease Control and Prevention (CDC) found that, between 12 February and 2 April 2020, children accounted for less than 2% of all cases of COVID-19 while making up 22% of the U.S. population (CDC COVID-19 Response Team 2020). A meta-analysis examining 16 studies as of 16 May 2020 found a pooled odds ratio of 0.44 of infections in children compared to adults (Viner et al. 2020a), while a contemporaneous study from the United Kingdom found no age-related infection differences (Woodhill 2020). A brief reopening of schools in Israel was followed by immediate closures two weeks later when safety guidelines were not strictly enforced during a heat wave (Stein-Zamir 2020). Children were also seen to travel to school via a range of transportation and to participate in extracurricular activities with differing groups of children.

Without traditional, full-day in-person school, children face food insecurity, slowed educational progression, and developmental harms, while mothers are disproportionately burdened with balancing work responsibilities with childcare (Esposito and Principi 2020, Viner et al. 2020b, Kneale et al. 2020). There is general agreement that schools should implement measures such as social distancing of at least six feet, lower teacher-student ratios, mask wearing, and isolation and discharge policies for children who become ill while at school (Riley 2020). These mitigations may not be fully feasible in terms of both staff and space for many school districts if all students are present. Many school districts are advocating for timely reopening to alleviate public health, economic, and emotional consequences for children and parents.

Several U.S. states have published and proposed reopening schedules for the 2020-2021 academic year, which typically begins in mid-August to early September. Many school districts and state Departments of Education are in favor of a “hybrid” or “blended rotation,” whereby one cohort of students are present on campus receiving face-to-face instruction, while another cohort or cohorts participates in distance learning (Hawai’i Department of Education 2020a, MCH Strategic Data 2020). Many schools also propose tiered plans, with fully in-person schooling under pandemic precautions of social distancing and mask usage eventually replacing fully online schedules once certain criteria have been met. Although the majority of states publicly share their school reopening plans (MCH Strategic Data 2020), few give any references to support their particular reopening decisions and whether they have considered other possible plans. As such, a need exists for a comparison between proposed school reopening plans to identify those with the potential to allow safe school reopenings.

We examined each U.S. state’s published school reopening plan to identify commonalities. Plans can be categorized by their proposed daily school capacity, division of school population into two or more, non-mixing cohorts, and by number of consecutive days spent in school. Examples of reopening plans include traditional five-day per week attendance at 100% school capacity, two-cohort half-capacity plans with students alternating days in attendance, and one day per grade 20% capacity plans (Bailey et al. 2020, MCH Strategic Data 2020). The majority of schools favor a “hybrid” or “blended rotation,” where one group of students are present on campus receiving face-to-face instruction while the other group(s) participates in distance learning (Hawai’i State Dept. of Ed 2020b, Illinois State Board of Ed 2020).

Policymakers are looking to predictive modeling efforts to understand the potential impacts of school reopening in the absence of extensive real-world data, but research to date is limited. Di Domenico et al. (2020) used demographic data from three regions in France coupled with a stochastic age-structured epidemic model to predict school closures in the absence of additional mitigation efforts in the wider community, such as telework, would have limited effect in reducing the magnitude of peak incidence.

This study aims to quantify the epidemic-level potential for several of the most common school reopening scenarios being considered in the United States to identify effective characteristics of each of these strategies to provide recommendations. We examine an idealized 100,000-person community with full susceptibility to COVID-19 infection, including 15,000 school-aged children. In all reopening scenarios, a single susceptible individual in the population is replaced with a clinically infected case.

## I. The Basic Model – No School

A single-patch deterministic model is initially postulated that captures all known methods of transmission by disease status as well as different pathways to testing. The population is stratified into nine epidemiologic compartments including Susceptible *S*, Exposed who will become subclinically infected *E*_*s*_, Exposed who will become clinically infected *E*, subclinically infectious *I*_*s*_, pre-clinically Infectious *I*_*p*_, clinically Infectious *I*_*c*_, Testing *K*, Quarantine *Q*, and Recovered *R*. It is assumed that all individuals are initially susceptible, with a single introduced infectious initiating an epidemic scenario. Susceptible individuals will contract infection at rate *β* times the proportion of infected individuals and then themselves become subclinically or clinically infectious, first passing through their appropriate exposed compartment at rate *ε*. Those who will become clinically infectious next pass through a preclinical compartment *I*_*P*_, where they have reduced infectiousness compared to clinically infectious cases *I*_*C*_. Infectious individuals, regardless of symptomology, recover from infection at rate *γ*.

Per understanding of COVID-19 disease transmission as of mid-August 2020, three routes of transmission are possible: from clinically infectious cases, from subclinically infectious cases at a potentially reduced rate, and from preclinically infectious cases at a known reduced rate. This population proportion of infectious individuals is summarized in the model as transmission parameter *B* = *β*(*I*_*C*_ + *σI*_*P*_ + *κI*_*S*_)/*N*. It is further assumed that a proportion *p* of clinically infectious individuals voluntarily seek confirmatory testing at rate *r*_1_ after symptom onset. While awaiting their positive test results, these individuals move to quarantine, assumed to be fully effective. For this study, any testing requirements to be released from quarantine are ignored and a 14-day quarantine period is assumed. Under mass testing, all non-quarantine compartments are tested with equal probability *q* at rate *r*_2_. Only preclinical, subclinical, and clinically infectious cases will have altered disease classification after testing; all other individuals will return to their current status compartment after testing. Figure 1 depicts the full hypothesized single-patch disease transmission model.

**FIGURE 1:**
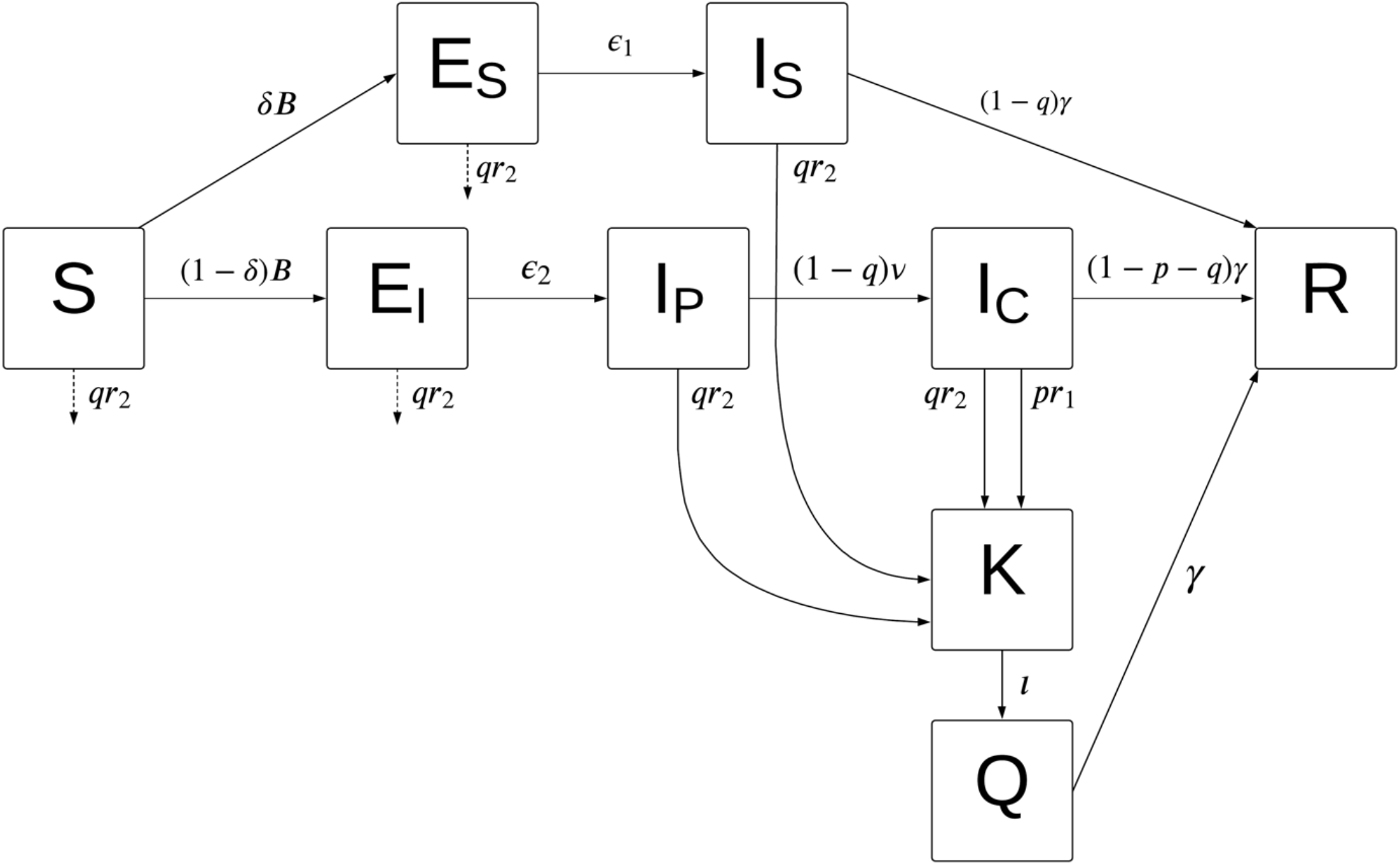
Simple single-patch transmission model. Diseased individuals move from susceptible *S* to exposed, *E*_*S*_ or *E*_*I*_ depending on whether the person ultimately expresses clinical symptoms or remains subclinical. Exposed individuals who will become clinically infectious next move to a preclinical phase, where they are infectious at a reduced rate *σ*. Subclinical infections *I*_*S*_ and clinical infections *I*_*C*_ recover to compartment *R* at rate *γ* if they do not receive positive test results. A proportion *p* of clinically infectious individuals seek voluntary COVID testing within 1/*r*_1_ days of symptom onset, with positive test results moving the person into quarantine *Q*. If mass testing begins for a proportion *q* of individuals, non-infectious individuals will not change disease compartments. Subclinically infectious cases and preclinically infectious cases will move to quarantine upon receipt of a positive test result. Quarantine is assumed to last 14 days, with serial negative testing requirements ignored for this study.

### Methods

To determine transmission rate *β*, a simplified single patch model that omits testing and quarantine was evaluated (Figure 2) to define the system’s basic reproductive number *R*_0_ based on model parameters (e.g., van den Driessche P, Watmough J 2002). This modified disease transmission model corresponds to the following set of ordinary differential equations:

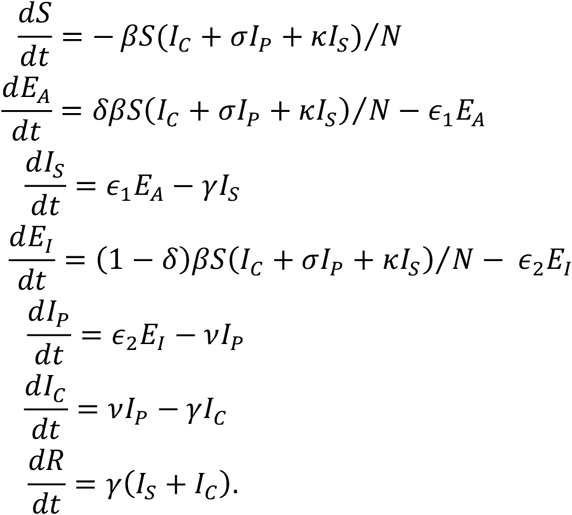

**FIGURE 2:**
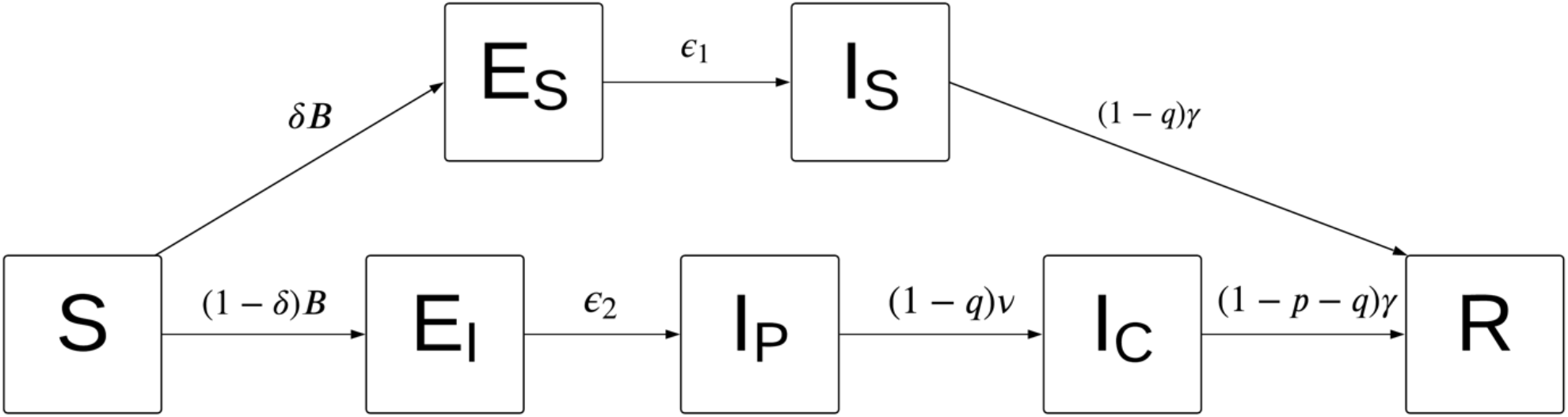
Simple single-patch transmission model at initial outbreak. The basic reproductive ratio *R*_0_ is defined as the expected number of secondary cases arising from an introduced primary case in an immunologically naïve population. When COVID-19 first appeared, there was no testing or quarantine acting to remove clinical infections *I*_*C*_ or subclinical infections *I*_*S*_ from the general public. This reduced model was used to define *R*_0_ by model parameter values to determine the transmission rate *β*.

At the beginning of an outbreak, when testing and case quarantine have not yet begun, *R*_0_ is a weighted average of subclinical, preclinical, and clinical contributions to secondary infections:

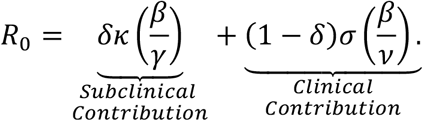

This equation for *R*_0_ can be algebraically solved for *β* and parameter values can be substituted from the “Best Guess” scenario outlined in the Centers for Disease Control and Prevention (CDC) “COVID-19 Pandemic Planning Scenarios” (CDC 2020). Reasonable probability distributions for each parameter were generated to reflect uncertainty in each value, as summarized in Table 1. Recovery time *γ* was not provided by the CDC and instead was taken as 10-14 days from preclinical onset based on local case observations. Parameter distributions and a uniformly distributed *R*_0_ ∼ *Unif*(2.5, 3.0) before interventions were implemented were used to determine a baseline point value and distribution for *β*, which conformed to a Gamma distribution with shape *α* = 15.45, rate *β* = 19.14 and expected value 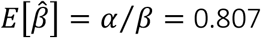. A second distribution under a reduced control reproductive number *R*_*C*_ ∼ *Unif*(1.01 − 1.25) was found to be *β*∼ *Gamma*(α = 14.03, *β* = 39.61), 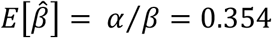.

**Table 1.** Descriptions and Distributions of Model Parameters.

| Parameter | Description | Expected Value | Distribution |
| --- | --- | --- | --- |
| $\delta$ | Proportion of infections that are subclinical | 0.4 | $\sim N(0.4, 0.075)$ |
| $B$ | Transmission rate, $= (I_C + \sigma I_P + \kappa I_A)/N$ | | |
| $\sigma$ | Reduced rate of preclinical infectiousness compared to fully infectious $I_C$ class | 0.35 | $\sim N(0.35, 0.05)$ |
| $\kappa$ | Reduced rate of subclinical infectiousness compared to fully infectious $I_C$ class | 0.7 | $\sim N(0.7, 0.1)$ |
| $1/\epsilon_1$ | Length of latent subclinical infection | 5 days | $\epsilon_1 \sim N(0.2, 0.01)$ |
| $1/\epsilon_2$ | Length of latent clinical infection | 4 days | $\epsilon_2 \sim N(0.25, 0.01)$ |
| $1/\nu$ | Length of preclinical infection | 1 day | $\nu \sim N(1, 0.01)$ |
| $1/\gamma$ | Length of COVID-19 infection | 8 days | $\gamma \sim N(1/8, 0.01)$ |
| $p$ | Proportion of clinically infected individuals who seek voluntary testing | 0.4 | $\sim Unif[0.0, 1.0]$ |
| $q_1$ | Proportion of “town” individuals tested under mass testing | 0.0 | $\sim Unif[0.0, 1.0]$ |
| $q_i$ | Proportion of “school” individuals tested under mass testing, $i$ = number of cohorts in plan | 0.0 | $\sim Unif[0.0, 1.0]$ |
| $1/r_1$ | Rate of time to seek testing | 4 days | |
| $1/r_2$ | Rate of mass testing | 7 days | |
| $R_0$ | Basic reproductive number at initial epidemic outbreak with no mitigations in place | 2.75 | $\sim Unif(2.5, 3.0)$ |
| $R_C$ | Control reproductive number under mitigations | 1.21 | $\sim Unif(1.01, 1.40)$ |
| $\beta_0$ | Transmission rate under initial outbreak conditions (i.e., calculated from $R_0$) | 0.807 | $\sim Gamma(15.28, 18.93)$ |
| $\beta_C$ | Transmission rate under mitigated conditions (i.e., calculated from $R_C$) | 0.354 | $\sim Gamma(14.03, 39.61)$ |

The full simple single-patch model with testing and quarantine was coded into R (version 4.0.1, “See Things Now”, R Core Team 2019) and evaluated over ten weeks as a baseline “summer vacation” scenario. To understand the potential behavior of the baseline model, a total of 10,000 simulations were run using parameters drawn from probability distributions as described in Table 1. Mean curves of susceptibles and infected, as well as confidence bands, are shown in Figure 3.

**FIGURE 3:**
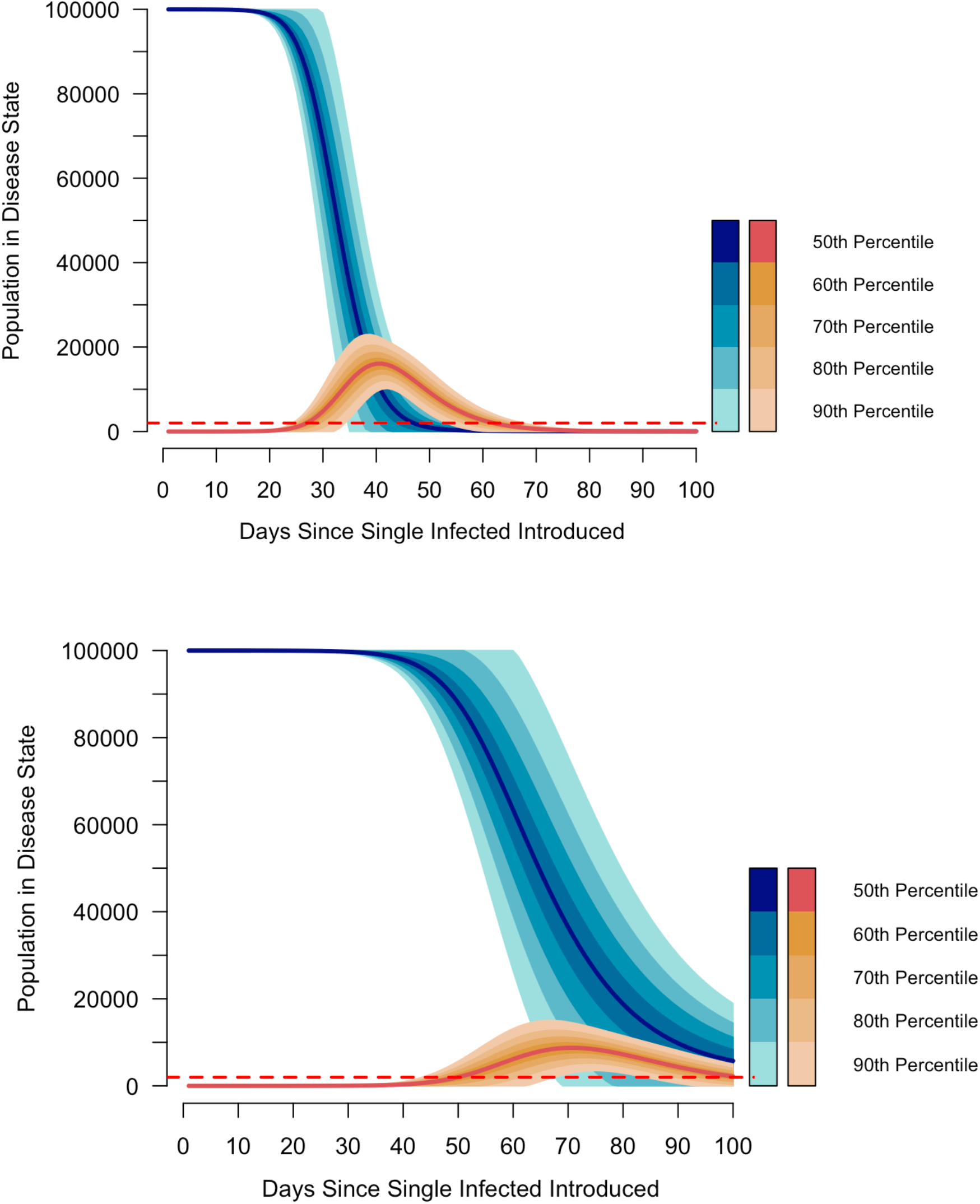
Full distance learning schedule. Under baseline conditions, an epidemic of more than 2,000 clinical and subclinical infections occurs on Day 27 and ends on Day 61, with 60.1% of the community infected. Under controlled conditions, the epidemic onset is delayed to Day 49 and ends on Day 100, with 59% of the community infected. This “flattening of the curve” does not reduce overall cases, but rather spreads them out through time to allow for the action of contact tracing and more restrictive mitigation measures to reduce community spread and to not overwhelm healthcare facilities.

### Results

In the baseline model under initial conditions with no schooling, a total of 60,089.49 (95% CI: 47,355.81, 72,823.17) clinical infections occur in a community of 100,000 individuals (Figure 3, upper). Since this model does not differentiate between schoolchildren and members of the surrounding community, this 60% infection prevalence includes 60.46% overall infection rate in children (9,068.94 cases, 95% CI: 5,725.98, 12,411.90). Mitigation levels that reduce *R*_*0*_ from 2.5 - 3.0 to just over 1.0 - 1.4, acts to “flatten the curve” without reducing total number of infections (Figure 3, lower).

## II. School Reopening Plans

School reopening plans divide the population into subgroups of “school” and “town” using parallel single-patch deterministic models so that individuals remain within their subgroup throughout disease progression. Such stratification is required when the homogeneous mixing assumption is expected to be violated, such as when children attend school and aren’t expected to be able to maintain the same mitigation levels to reduce transmission as with adults who do not attend school. The added model complexity can allow for emergent dynamics when a different set of parameters pertain to a subset of the population.

In the simplest school reopening model, depicted in figure 4(a), a percentage of the population is deemed the “school” population. Interaction between subgroups is specified via a transmission matrix that modifies the transmission rate *β* calculated for the base model depending on type of interaction: student-student interactions are expected to have a transmission rate > *β* while in school due to a higher frequency of interactions, while student-town interactions are expected to have a transmission rate < *β* to represent reduced interactions with adults to only parents and school staff during school days. Under this simplest school reopening scenario, we assume a student-student transmission rate that is 20% higher than that seen between non-school individuals, an 80% reduced transmission rate between students and town during school days, and a 60% reduced transmission between students and town during distance learning days (if applicable to the plan):

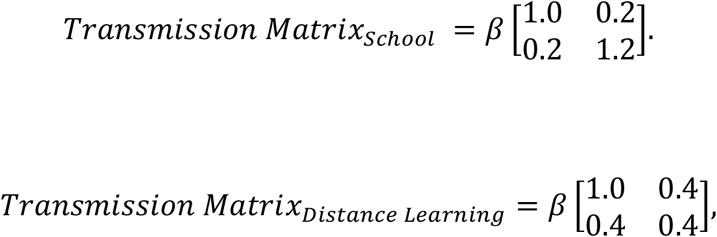

with row corresponding to Town, School and columns corresponding to Town, School.

**FIGURE 4.**
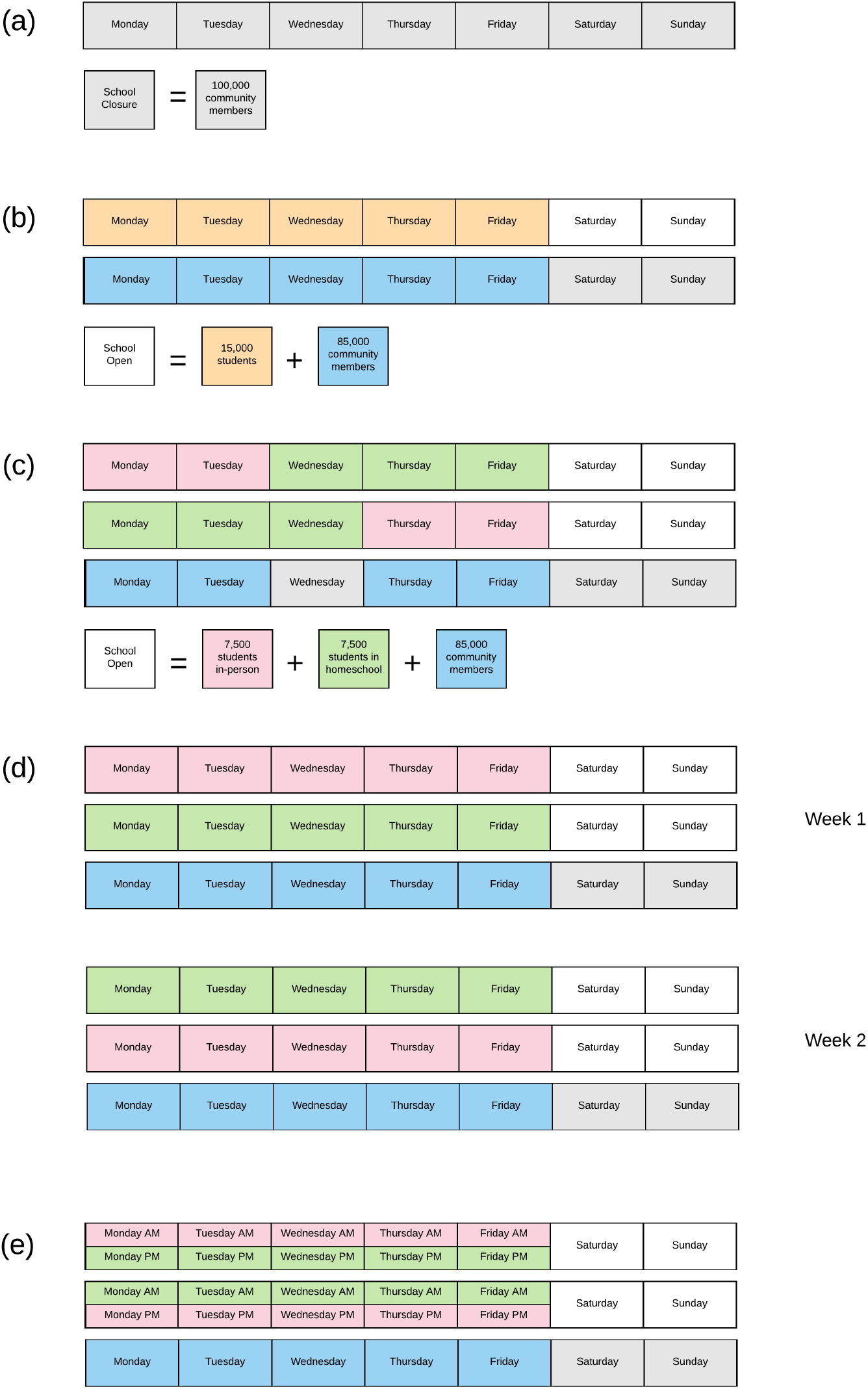
School Reopening Plans. In baseline (a), no children attend in-person school. This is equivalent to summer vacation, with all community members interacting and transmitting disease at rate *β*_0_ under uncontrolled conditions and *β*_*C*_ under controlled conditions. In (b), school is open five days per week at 100% capacity, or 15,000 students, reducing the community to 85,000 intermixing non-school individuals. For reduced full day scenarios, some school days are replaced with distance learning mixing conditions on some days. In the two-day switch model (c), students are split into two, non-mixing cohorts with a first cohort of 7,500 students attending school for two days, followed by one distance learning day for facility cleaning, and then the next 7,500-student cohort attends for two days. In the five-day switch model (d), a first 7,500-student cohort attends for five consecutive days while the second 7,500-student cohort is distance learning at a reduced community mixing rate. In the second week, the first cohort is distance learning while the second 7,500-student cohort attends in-person schooling. In the AB school reopening plan (e), two 7,500-student cohorts attend school every day, with one group attending in the morning and the other attending in the afternoon.

This simplest school reopening model is evaluated for a traditional five-day week, as well as in reduced scenarios of four days per week, three days per week, and one day per week under both an *R*_*0*_ of 2.5 - 3.0 without mitigations and under an *R*_*C*_ of 1.01 - 1.40 with mitigations. As a baseline comparison, a full distance learning plan, i.e., zero days per week attendance, is examined under both uncontrolled and controlled transmission.

### Methods

The baseline 0% capacity school function in R was adjusted to a two-subgroup model with a larger “town” and smaller “school” group. Susceptible individuals are exposed to infected individuals in town and school using a transmission matrix. The full student body of 15,000 students (15% of the 100,000-person population) attends school for the specified number of days, which is modeled as the two subgroups running through compartmentalized disease states in parallel. A complete week under a school reopening scenario consists of *days in school + days in distance learning + days in weekend + 1 day*, where each change in population-level interaction is modeled using the base mixing model run with the appropriate transmission matrix and parameters drawn from the stochastic parameter ranges described in Table 1, and for time steps of *days + 1*, where the extra day is used to define initial compartmental conditions for the next week. Each single week school schedule function was run through ten weeks. The total number of clinically infectious cases, total number of clinical infections in children, and percentage of children with clinical infections were retained for each model.

Two scenarios were examined for each single cohort school schedule: Scenario A refers to an initially infected individual introduced into the town subgroup and Scenario B refers to an initially infected individual occurring in the school subgroup. A baseline of zero days per week, full distance learning model was also run for comparison.

### Results

Table 3 reports the findings for each school scenario run under initial outbreak conditions and Table 4 reports findings under mitigated conditions. Increased consecutive days in school offers a slight reduction in total number of cases compared to the baseline of no schooling by delaying exposure between cohorts. Differences between Scenario A and Scenario B are negligible. Figure 4 shows mean total cases under Scenario A under *R*_*0*_-type transmission rates and under *R*_*C*_-type transmission rates. Mean total cases between the two transmission rates are also nearly identical, but confidence limits under each transmission condition vary and showing differing trends. Under *R*_*0*_ conditions, confidence intervals show a bowtie pattern, narrowing from baseline to three consecutive school days, then widening again out to five consecutive school days. Under *R*_*C*_ conditions, confidence intervals are overall lower than for *R*_*0*_ conditions and are skewed compared to the mean, indicating a probability distribution of means which more often includes very small outbreaks.

**Table 2.** 100% School Capacity Scenarios Under *R*_*0*_, Reduced Mixing on Distance Learning Days, and Regular Community Mixing on Off Days.

| Plan | Total Cases<br>(95% CI) | Cases in Children<br>(95% CI) | % Cases in Children<br>(95% CI) |
| --- | --- | --- | --- |
| **Scenario A: Infected individual introduced into the community** |  |  |  |
| BASELINE 1 = No School | 60,089.49 (47,355.81, 72,823.17) | 9,068.94 (5,725.98, 12,411.90) | 60.46% (38.17, 82.75) |
| BASELINE 2 = Full Distance learning | 59,942.76 (50,767.00, 69,118.52) | 8,958.00 (6,808.34, 11,107.66) | 59.72% (45.39, 74.05) |
| One Day Per Week | 58,943.63 (50,842.77, 67,044.50) | 8,079.44 (6,210.02, 9,948.85) | 53.86% (41.40, 66.33) |
| Two Days Per Week<br>T | 58,154.25 (50,863.98, 65,444.53) | 7,271.25 (5,580.71, 8,961.80) | 48.48% (37.20, 59.75) |
| Three Days Per Week | 57,424.01 (50,349.01, 64,499.02) | 6,521.76 (4,914.96, 8,128.55) | 43.48% (32.77, 54.19) |
| Four Days Per Week | 56,742.52 (49,209.45, 64,275.60) | 5,869.49 (4,224.66, 7,514.32) | 39.13% (28.16, 50.10) |
| Five Days Per Week | 56,009.53 (47,295.70, 64,723.36) | 5,263.53 (3,565.85, 6,961.20) | 35.09% (23.77, 46.41) |
| **Scenario B: Infected individual introduced into the school** |  |  |  |
| BASELINE 2 = Full Distance learning | 59,846.55 (50,541.50, 69,151.59) | 8,951.37 (6,792.88, 11,109.86) | 59.68% (45.29, 74.07) |
| One Day Per Week | 59,063.94 (50,933.64, 67,194.23) | 8,085.73 (6,224.85, 9,946.61) | 53.90% (41.50, 66.31) |
| Two Days Per Week<br>T | 58,109.36 (50,828.89, 65,389.83) | 7,255.18 (5,555.28, 8,955.09) | 48.37% (37.04, 59.70) |
| Three Days Per Week | 57,452.02 (50,351.76, 64,552.27) | 6,524.49 (4,900.71, 8,148.27) | 43.50% (32.67, 54.32) |
| Four Days Per Week | 56,812.03 (49,263.95, 64,360.11) | 5,865.66 (4,228.33, 7,503.00) | 39.10% (28.19, 50.02) |
| Five Days Per Week | 56,035.13 (47,339.54, 64,730.71) | 5,262.56 (3,559.09, 6,966.04) | 35.08% (23.73, 46.44) |

**Table 4.**
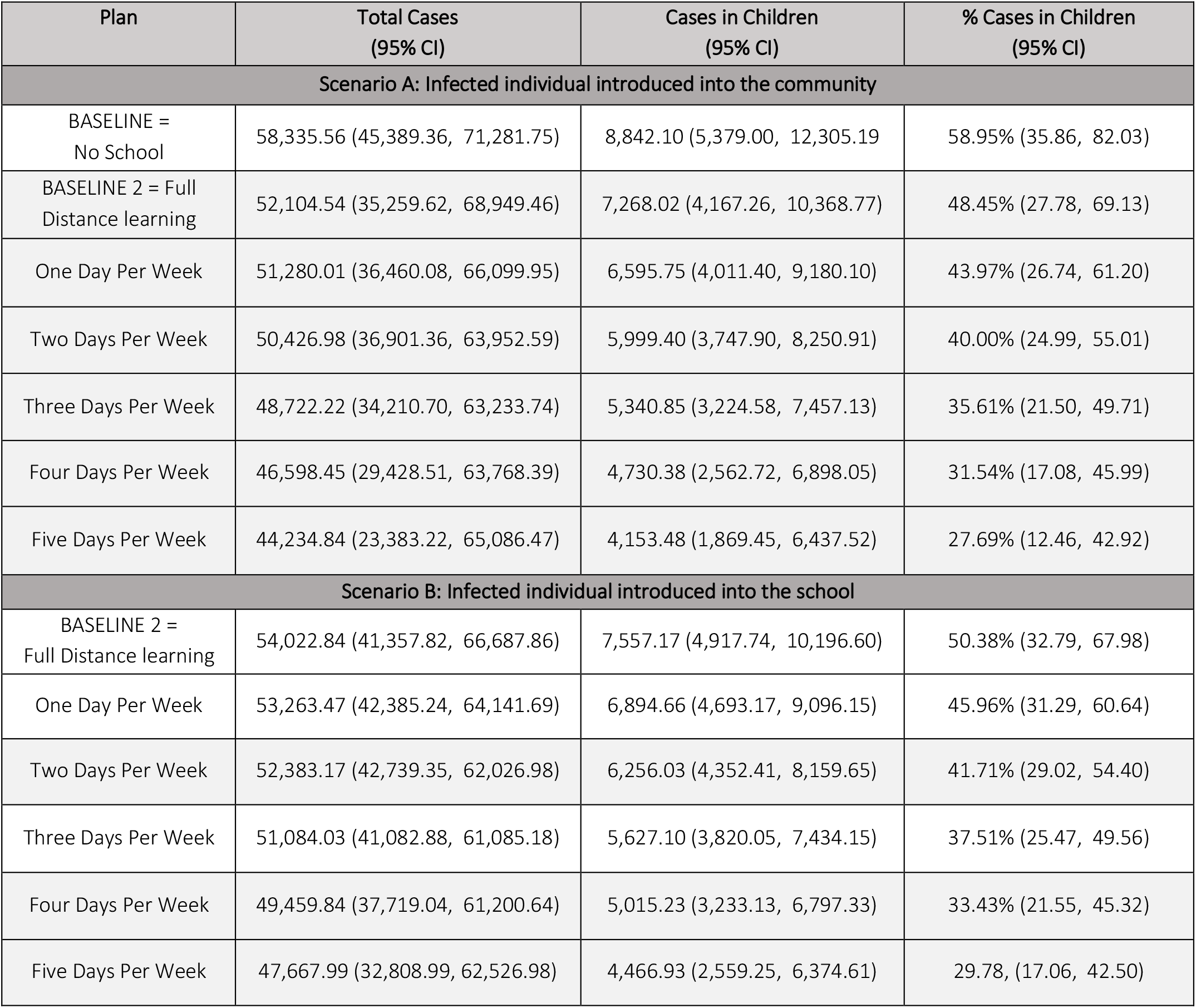
100% Reopening Scenarios Under *R*_*C*_, Reduced Mixing on Distance learning Days, and Regular Community Mixing on Off Days.

| Plan | Total Cases<br>(95% CI) | Cases in Children<br>(95% CI) | % Cases in Children<br>(95% CI) |
| --- | --- | --- | --- |
| **Scenario A: Infected individual introduced into the community** |  |  |  |
| BASELINE =<br>No School | 58,335.56 (45,389.36, 71,281.75) | 8,842.10 (5,379.00, 12,305.19) | 58.95% (35.86, 82.03) |
| BASELINE 2 = Full<br>Distance learning | 52,104.54 (35,259.62, 68,949.46) | 7,268.02 (4,167.26, 10,368.77) | 48.45% (27.78, 69.13) |
| One Day Per Week | 51,280.01 (36,460.08, 66,099.95) | 6,595.75 (4,011.40, 9,180.10) | 43.97% (26.74, 61.20) |
| Two Days Per Week | 50,426.98 (36,901.36, 63,952.59) | 5,999.40 (3,747.90, 8,250.91) | 40.00% (24.99, 55.01) |
| Three Days Per Week | 48,722.22 (34,210.70, 63,233.74) | 5,340.85 (3,224.58, 7,457.13) | 35.61% (21.50, 49.71) |
| Four Days Per Week | 46,598.45 (29,428.51, 63,768.39) | 4,730.38 (2,562.72, 6,898.05) | 31.54% (17.08, 45.99) |
| Five Days Per Week | 44,234.84 (23,383.22, 65,086.47) | 4,153.48 (1,869.45, 6,437.52) | 27.69% (12.46, 42.92) |
| **Scenario B: Infected individual introduced into the school** |  |  |  |
| BASELINE 2 =<br>Full Distance learning | 54,022.84 (41,357.82, 66,687.86) | 7,557.17 (4,917.74, 10,196.60) | 50.38% (32.79, 67.98) |
| One Day Per Week | 53,263.47 (42,385.24, 64,141.69) | 6,894.66 (4,693.17, 9,096.15) | 45.96% (31.29, 60.64) |
| Two Days Per Week | 52,383.17 (42,739.35, 62,026.98) | 6,256.03 (4,352.41, 8,159.65) | 41.71% (29.02, 54.40) |
| Three Days Per Week | 51,084.03 (41,082.88, 61,085.18) | 5,627.10 (3,820.05, 7,434.15) | 37.51% (25.47, 49.56) |
| Four Days Per Week | 49,459.84 (37,719.04, 61,200.64) | 5,015.23 (3,233.13, 6,797.33) | 33.43% (21.55, 45.32) |
| Five Days Per Week | 47,667.99 (32,808.99, 62,526.98) | 4,466.93 (2,559.25, 6,374.61) | 29.78, (17.06, 42.50) |

**Table 5:**
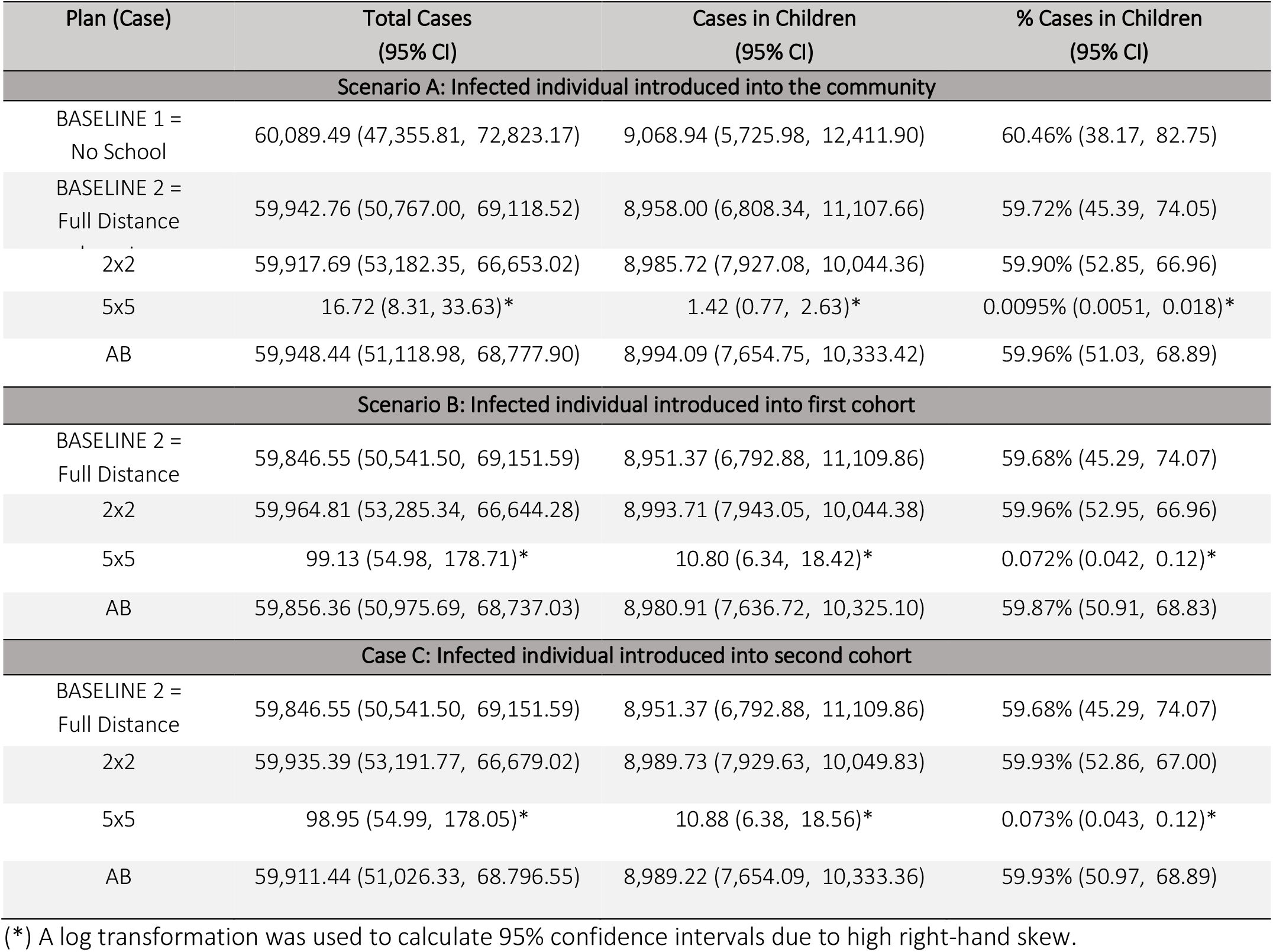
50% School Scenarios Under *R*_*0*_ with reduced school-community mixing on distance learning days.

**Table 6:** 50% School Scenarios Under *R*_*C*_ with reduced school-community mixing on distance learning days. To aid comparisons, the baseline no-school scenario under *R*_*C*_ conditions is provided in the top row.

| Plan (Case) | Total Cases<br>(95% CI) | Cases in Children<br>(95% CI) | % Cases in Children (95% CI) |
| --- | --- | --- | --- |
| **Scenario A: Infected individual introduced into the community** |  |  |  |
| BASELINE 1 =<br>No School | 58,335.56 (45,389.36, 71,281.75) | 8,842.10 (5,379.00, 12,305.19) | 58.95% (35.86, 82.03) |
| BASELINE 2 =<br>Full Distance | 52,104.54 (35,259.62, 68,949.46) | 7,268.02 (4,167.26, 10,368.77) | 48.45% (27.78, 69.13) |
| 2x2 | 95.77 (47.14, 194.55)* | 11.85 (6.04, 23.28) | 0.079% (0.040, 0.16) |
| 5x5 | 3.42 (2.17, 5.41)* | 0.19 (0.12, 0.33)* | 0.0013% (0.00077, 0.0022)* |
| AB | 17,541.99 (3,241.76, 43,245.67)† | 2,812.03 (561.14, 6,784.60)† | 18.75% (3.74, 45.23)† |
| **Scenario B: Infected individual introduced into first cohort** |  |  |  |
| BASELINE 2 =<br>Full Distance | 59,846.55 (50,541.50, 69,151.59) | 8,951.37 (6,792.88, 11,109.86) | 59.68% (45.29, 74.07) |
| 2x2 | 851.28 (436.29, 1,661.02)* | 109.64 (56.54, 212.62)* | 0.73% (0.38, 1.42)* |
| 5x5 | 15.06 (9.20, 24.66)* | 1.71 (1.06, 2.77)* | 0.11% (0.0070, 0.018)* |
| AB | 46,836.11 (25,592.40, 61,092.31)‡ | 7,196.86 (4,056.68, 9,334.50)‡ | 47.98% (27.04, 62.23)‡ |
| **Scenario C: Infected individual introduced into second cohort** |  |  |  |
| BASELINE 2 =<br>Full Distance | 59,846.55 (50,541.50, 69,151.59) | 8,951.37 (6,792.88, 11,109.86) | 59.68% (45.29, 74.07) |
| 2x2 | 765.48 (393.84, 1,487.82)* | 98.32 (51.11, 189.16)* | 0.66% (0.34, 1.26)* |
| 5x5 | 14.99 (9.14, 24.58)* | 1.74 (1.08, 2.83)* | 0.012% (0.0072, 0.019)* |
| AB | 45,627.66 (27,475.22, 63,780.09) | 7,013.37 (4,293.67, 9,733.07) | 46.76% (28.62, 64.89) |

(\*) A log transformation was used to calculate 95% confidence intervals due to high right-hand skew.
(†) A square root transformation was used to calculate the 95% confidence interval due to right-hand skew.
(‡) A squaring transformation was used to calculate the 95% confidence interval due to left-hand skew.

**Table 7:** 20% Cohort Reopening Plan Under *R*_*0*_.

| Plan (Case) | Total Cases<br>(95% CI) | Cases in Children<br>(95% CI) | % Cases in Children<br>(95% CI) |
| --- | --- | --- | --- |
| BASELINE 1 =<br>No School | 60,089.49 (47,355.81, 72,823.17) | 9,068.94 (5,725.98, 12,411.90) | 60.46% (38.17, 82.75) |
| BASELINE 2 =<br>Full Distance learning | 59,942.76 (50,767.00, 69,118.52) | 8,958.00 (6,808.34, 11,107.66) | 59.72% (45.39, 74.05) |
| One Day Per Grade<br>(Infectious in Town) | 176.90 (48.02, 651.68)* | 26.33 (7.08, 97.94)* | 0.18% (0.047, 0.65)* |
| One Day<br>(Infectious in Day 1) | 981.04 (329.38, 2,921.98)* | 146.96 (49.28, 438.31)* | 0.98% (0.33, 2.92)* |
| One Day<br>(Infectious in Day 2) | 981.53 (333.11, 2,892.18)* | 147.04 (49.83, 433.83)* | 0.98% (0.33, 2.89)* |
| One Day<br>(Infectious in Day 3) | 972.61 (326.93, 2,893.46)* | 145.70 (48.91, 434.03)* | 0.97% (0.33, 2.89)* |
| One Day<br>(Infectious in Day 4) | 972.40 (329.34, 2,871.07)* | 145.67 (49.27, 430.68)* | 0.97% (0.33, 2.87)* |
| One Day<br>(Infectious in Day 5) | 981.69 (335.62, 2,871.47)* | 147.06 (50.21, 430.72)* | 0.98% (0.33, 2.87)* |

(\*) A log transformation was used to calculate 95% confidence intervals due to high right-hand skew.

**Table 8:** 20% Cohort Reopening Plan under *R*_*C*_.

| Plan (Case) | Total Cases<br>(95% CI) | Cases in Children<br>(95% CI) | % Cases in Children<br>(95% CI) |
| --- | --- | --- | --- |
| BASELINE 1 =<br>No School | 58,335.56 (45,389.36, 71,281.75) | 8,842.10 (5,379.00, 12,305.19) | 58.95% (35.86, 82.03) |
| BASELINE 2 =<br>Full Distance learning | 52,104.54 (35,259.62, 68,949.46) | 7,268.02 (4,167.26, 10,368.77) | 48.45% (27.78, 69.13) |
| One Day Per Grade<br>(Infectious in Town) | 10.13 (4.22, 24.36)* | 1.34 (0.50, 3.60)* | 0.0090% (0.0033, 0.024)* |
| One Day<br>(Infectious in Day 1) | 51.64 (23.24, 114.77)* | 7.58 (3.35, 17.14)* | 0.051% (0.022, 0.11)* |
| One Day<br>(Infectious in Day 2) | 51.51 (23.13, 114.72)* | 7.56 (3.33, 17.13)* | 0.050% (0.022, 0.11)* |
| One Day<br>(Infectious in Day 3) | 51.17 (22.87, 114.48)* | 7.51 (3.30, 17.09)* | 0.050% (0.022, 0.11)* |
| One Day<br>(Infectious in Day 4) | 51.81 (23.26, 115.38)* | 7.60 (3.35, 17.23)* | 0.051% (0.022, 0.11)* |
| One Day<br>(Infectious in Day 5) | 51.95 (23.22, 116.27)* | 7.62 (3.35, 17.37)* | 0.051% (0.022, 0.12)* |

(\*) A log transformation was used to calculate 95% confidence intervals due to high right-hand skew.

Figure 5 shows mean number of cases in children. In a model run for ten weeks with initial conditions of a single converted infective case in the community, children show a perceptible dose-response relationship to additional days in in-person schooling, reducing from approximately 9,069 infections under initial epidemic conditions without schooling to 5,264 cases, a 42% reduction, under full five-day schooling. Under controlled conditions, cases in children reduce from approximately 8,086 cases to 5,263, a 35% reduction.

**FIGURE 5:**
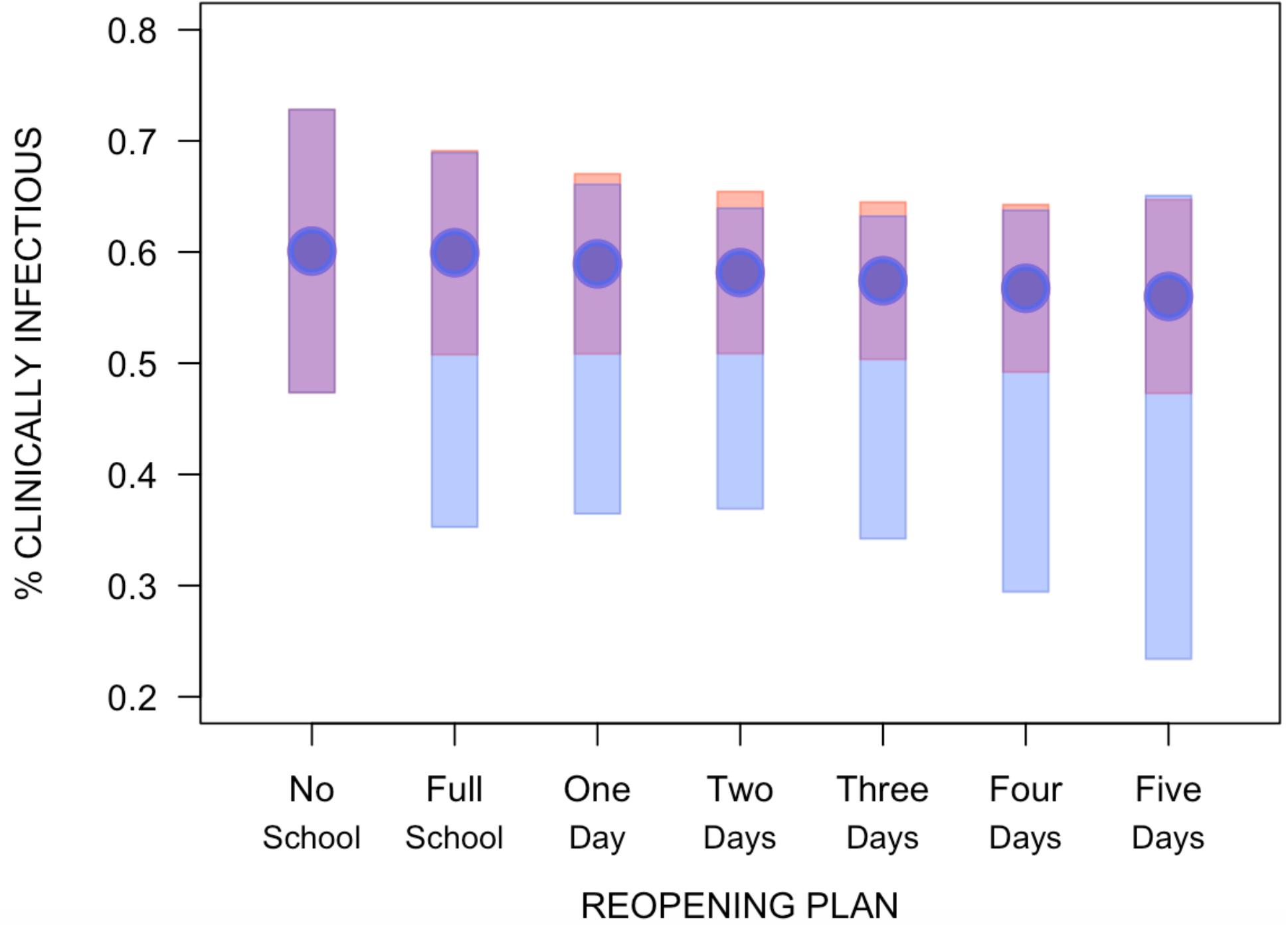
Proportion of Clinically Infectious Individuals Under Differing Transmission Rates in Single Cohort, 100% School Capacity Plans. Proportion of clinically infectious cases within community under *R*_*0*_ conditions (red dots) shows an overall downward trend and protective effect of each additional case of schooling compared to baseline, with a bowtie pattern of respective confidence intervals on these means, with initially narrower confidence intervals giving way to wider intervals after three days of consecutive schooling. Proportion of clinically infectious individuals under *R*_*C*_ conditions (blue dots) shows a nearly identical overall downward trend and protective effect of additional schooling when overplotted onto the proportions calculated under *R*_*0*_ conditions, as well as a bowtie pattern for confidence intervals. Confidence intervals for school plans under *R*_*C*_ do not as readily overlap with confidence intervals, indicating that the overall community transmission rate plays a large role in the variability seen between simulations.

**FIGURE 6:**
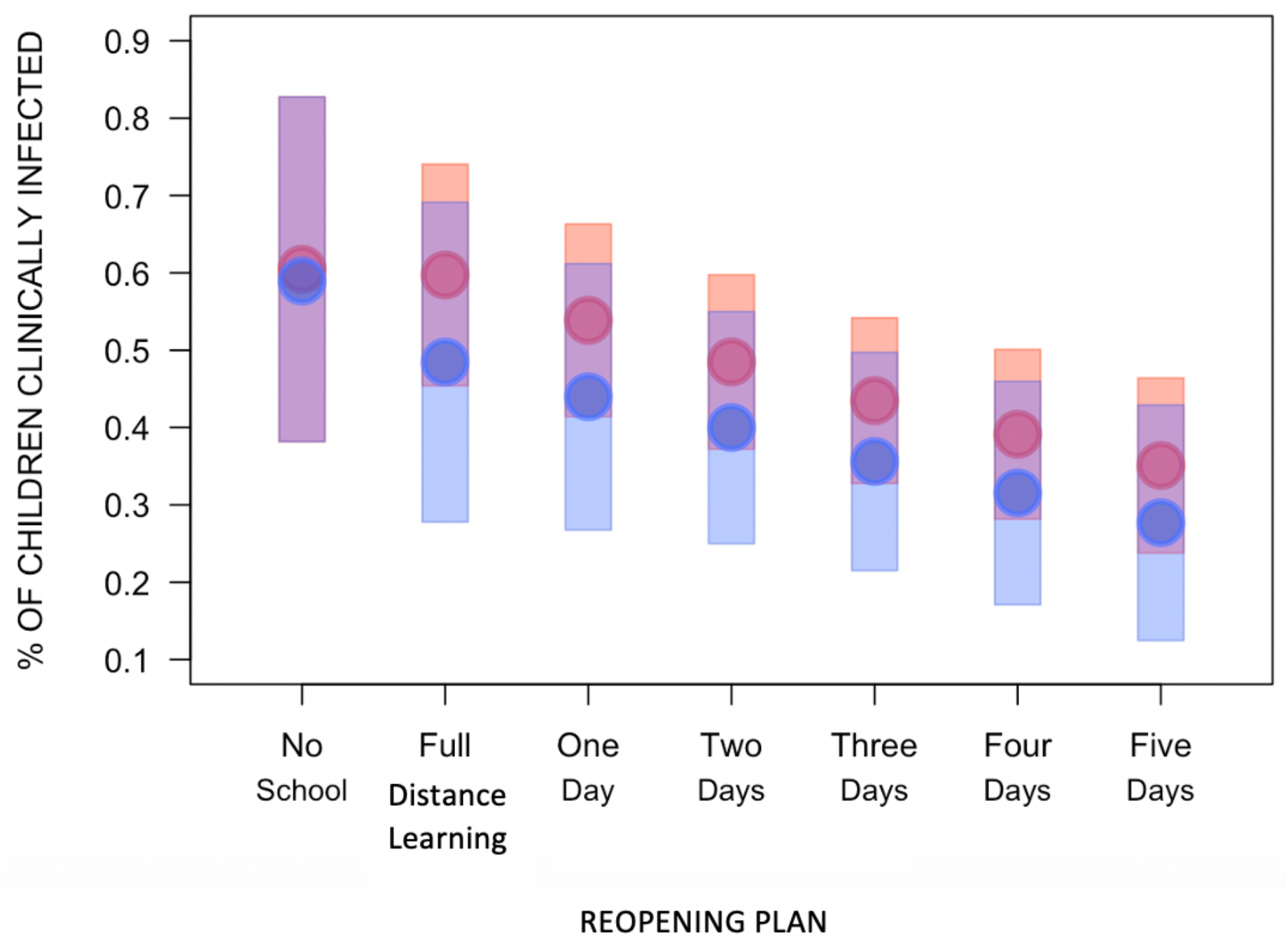
Proportion of Children with Clinical Infections by Consecutive Days in School Plans. A dose-response between number of days in school and reduced number of cases in children is seen under both uncontrolled transmission (red) and controlled transmission (blue), though these differences are not statistically significant, as demonstrated by their overlapping confidence intervals. After ten weeks of school reopening and with no change in transmission rates, there are less overall cases in children under controlled transmission levels.

### Discussion

Number of continuous days in school has a protective effect compared to no school by temporarily protecting subpopulations from active infection, as demonstrated by the five-day scenario being more protective than the one-day scenario. The increased interaction within school is seemingly offset by their highly reduced interaction with the larger community, which acts to isolate a significantly sized subpopulation. The non-overlapping confidence intervals between *R*_*0*_ conditions and *R*_*C*_ conditions indicates that further decreasing overall community spread will be critical for preventing cases within schools. School is most effective for the student population. Further reduction in childhood infectious cases is expected if the student cohort is additionally subdivided, as in several school reopening scenarios proposed by school districts. While these findings and trends are not statistically significant due to included noise in estimated parameters, further research and understanding of these parameters will reduce such noise and make the protective effect of days in school more pronounced.

## III. Two Cohort Reopening Plans

Under two-cohort school reopening plans, students are split into two or five cohorts. Like in single cohort scenarios, cohorts attend school together for consecutive days, with greatly reduced mixing between students and town assumed on in-person school days and less reduced mixing between students and town assumed on distance learning days. Cohorts intermix with one another at reduced rates in the same way that students interacted with town at reduced rates in the single cohort scenarios, but the additional school cohort requires an expansion of the transmission matrix. If the school reopening plan includes a cleaning day, this day is treated as a distance learning day for both cohorts. The transmission matrix for Cohort 1 on in-person schooling days is shown below:

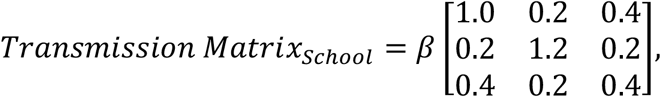

with rows corresponding to Town, Cohort 1, Cohort 2 and columns corresponding to Town, Cohort 1, Cohort 2.

The transmission matrix on cleaning days if applicable:

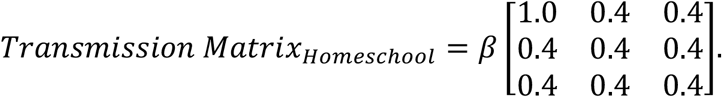

A final proposed 50% capacity, two-cohort plan studied is the AB plan. Under this plan, the two cohorts each attend school in person every day for a half school day, with one cohort attending in the morning and the other attending in the afternoon. We assume under this plan that students do not intermix on their way to and from school, and that classroom surfaces and other shared facilities cannot transmit COVID-19 between cohorts.

The transmission matrix used on all school days is:

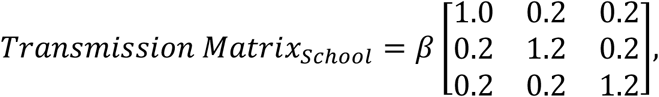

with rows and columns again corresponding to Town, Cohort 1, and Cohort 2, respectively.

### Methods

The single-cohort function in R is modified to add an additional school cohort, so that three cohorts interact. Functions were written to model a single week, as depicted in Figure 4(c-e). In the two-day switch model, shown in Figure 4(c), a first cohort of 50% of students attend in-person on Monday and Tuesday, then the school is closed for cleaning on Wednesday. The second cohort of students attend Thursday and Friday, then all students are home for a regular weekend on Saturday and Sunday. The full week is modeled as *two days of Cohort 1 in school/Cohort 2 in distance learning + one day of full distance learning + two days of Cohort 1 in distance learning/Cohort 2 in school + two days of weekend + 1 day*, with appropriate transmission matrices used for each run of the base mixing model. Once again, the week includes one additional day to generate initial conditions for the next week. A subsequent function then runs each scenario for ten weeks and retains total number of clinically infectious cases, number of clinically infectious cases in children, and percentage of children clinical infectious in each of 10,000 model simulations. Confidence intervals were calculated.

In the five-day cohort switching model, shown in Figure 4(d) and which takes two weeks to repeat, the model is run for five weeks instead of ten, omitting the cleaning day between cohorts. The full model, repeated for five runs to total ten weeks, is *five days of Cohort 1 in school/Cohort 2 in distance learning + two days of weekend + five days of Cohort 1 in distance learning/Cohort 2 in school + two days of weekend + 1 day*.

In the AB plan model, shown in Figure 4(e), the five-day 100% capacity model was used, with the transmission matrix expanded for school days to three cohorts in total. Since all students are in school for part of the day and in distance learning for the remainder of the day, only a single school-day transmission matrix is required:

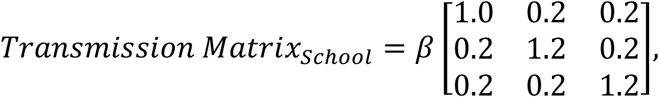

where rows and columns correspond to Town, Cohort 1, and Cohort 2, respectively and it is assumed that all student interactions occur as in a full school day. The single week model is *five days of Cohort 1/Cohort 2 in school and in distance learning + two weekend days + 1 day*.

### Results

The 50% capacity, two-day switch model under *R*_*0*_ conditions is not effective in reducing cases counts when compared to either of the baseline comparisons. This reopening plan is effective in reducing epidemic-level case counts under *R*_*C*_ conditions, however, though its effectiveness varies significantly depending on which scenario of placement of converted infectious case. The two-day plan is especially effective when the converted infective is a townsperson, as opposed to a student, reducing overall cases from 58,335.56 to 95.77 (95% CI: 47.14, 194.55) cases, a 99.8% reduction in cases.

Under both uncontrolled and controlled levels of transmission, the 50% capacity, five-day switch model is very effective in reducing overall case counts and case counts in children. Under both conditions, the five-day switch model is especially effective at reducing infection in children when the introduced infective is a townsperson, reducing overall cases from 60,089.49 cases in the baseline model to 16.72 cases (95% CI: 8.31, 33.63), a 99.97% reduction in cases in the uncontrolled scenario and from 58,335.56 cases to 3.42 (95% CI: 2.17, 5.41) cases, a reduction over 99.99%.

The AB school plan, which consists of two cohorts of students each alternately attending in-person school for a half-day and distancing learning for a half-day, is not effective in reducing overall cases or cases in children under uncontrolled transmission conditions. Under controlled transmission conditions, the AB school model is somewhat effective in reducing overall cases, especially when the initial infective is in town, reducing total infections from 52,104.54 total cases under baseline distance learning to 17,541.99 (95% CI: 3,241.76, 43,245.67) total cases under the AB reopening plan, a 66% reduction in infection. With regard to infections in children, the AB reopening plan reduced total cases from 7,268.02 cases under baseline distance learning to 2,812.03 (95% CI: 561.14, 6,784.60) cases, a 61% reduction in infection. When the introduced infective is in either school cohort, infection reduction is less effective, reducing total cases by approximately 10% and cases in children by less than 1%.

### Discussion

Split cohort plans in general are very robust against introduced infectious cases, especially when the initial infection occurs within the town rather than the school population. The AB plan is overall less effective than either the two-day or five-day switch plans, and may not be effective at all in uncontrolled transmission situations. It must be recalled that this modeling exercise assumes no current infections in the community at the start of school and that people continue to maintain control measures even in the absence of active community cases. This modeling exercise also assumes that students maintain reduced community mixing on distance learning days, which is more likely to occur when students receive live virtual lessons with required attendance.

## IV. Five Cohort Plan

In the five cohort reopening plan students form cohorts by grade level, with each grade level using school facilities for a single day of in-person instruction. This type of plan makes the most sense for private schools or public elementary schools, where students generally receive grade-specific instruction, and would likely be more difficult in high schools were students are less strictly divided by grade level.

### Methods

The two-cohort, 50% school capacity R function was further modified to allow a total of five interacting school cohorts. Once more, each school cohort, as well as the town, progress through disease states in parallel, only interacting via a transmission matrix specifying the levels of interactions between cohorts with respect to an overall rate. The cohort attending in-person school interacts with each other at 120% *β* and interacts with all other cohorts, including town, at a reduced 20% *β*. Distance learning students are expected to interact at a reduced rate with all other cohorts, including their own. The transmission matrix for Day is:

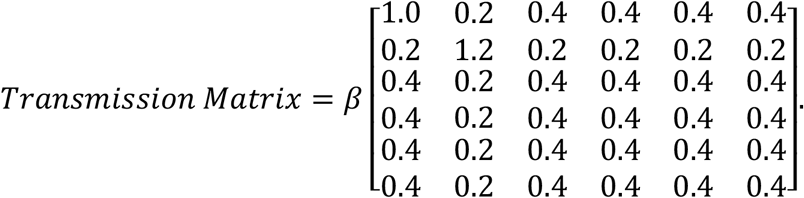

The non-school town interacts at rate *β*, with in-person schooling students at a reduced rate of 20% *β*, and with distance-learning students at a reduced rate of 40% *β*. A separate transmission matrix is used for each school day corresponding to the cohort that experiences in-person school, with all other students in distance learning. Finally, the weekend consists of an identity matrix multiplier on transmission for three days, with the third day serving as initial conditions for the next week. This model was run over ten consecutive weeks as well as six cases overall, one for each cohort with a single susceptible replaced with an infectious.

### Results

Under uncontrolled conditions, the 20% capacity, Five Cohort school reopening plan reduces total infections from 59,942.76 in the distance learning baseline to 176.90 (95% CI:48.02, 651.68, a 99.7% reduction) when the converted infectious individual is in town. Clinically infectious cases in children is reduced from 8,958.00 to 26.33 (95% CI: 7.08, 97.94) when the converted infectious individual is in town, also a 99.7% reduction in cases. When the converted infectious person is a child, total cases in town are reduced to roughly 978 cases, a 93.48% reduction from the full distance learning baseline.

Under controlled conditions, the 20% capacity, Five Cohort school reopening plan reduces total infections from 52,104.54 in the full distance learning baseline to 10.13 (95% CI: 4.22, 24.36) total cases, a greater than 99.99% reduction, when the converted infectious person is In town. When the converted infectious individual is a student, total cases are reduced to slightly less than 52, a greater than 99.95% reduction from baseline comparison.

### Discussion

The 20% capacity, Five-Cohort school reopening plan is very protective against a single converted infectious individual when the overall community has no other active cases, both when transmission is controlled through protective measures and when it is not. This reopening plan may be less feasible in practice, however, if a school is unable to operate with only 20% of students in-person per day. This type of reopening plan will probably be more realistic for elementary schools than middle schools or high schools, because students generally do not move between classrooms. If students attend classes

## V. CONCLUSIONS AND NEXT STEPS

All school reopening plans were examined under both non-controlled epidemic conditions and under controlled conditions. It is assumed that the overall population is initially disease-free and fully susceptible to COVID-19 before an initial infection occurs, either in the town or in schools. If these two groups regularly interact outside of school attendance, the location of the initial infection has little effect. A protective effect is seen with increased consecutive days in school compared to no school or distance learning, though the magnitude of this effect is small. School reopening plans that reduce a single large population into several sub-populations and limit their interaction throughout the week are the most effective strategies to reduce overall transmission. If schoolchildren are isolated at school during school hours but then move around freely within the larger community during non-school hours, or do not stay home during designated distance learning days, the protective effect of separating the population is will likely be lost. The most effective plan is the “Five-Day Switch Plan,” which takes advantage of both benefits by separating school children into two cohorts and using five consecutive school days per cohort.

In follow up to this study, we will perform a sensitivity analysis of the most promising scenarios to assess which parameters are most driving these effects. We will additionally examine threshold levels of transmission differences between school and town populations, and levels and timing of mass testing, to maintain very low levels of overall cases, as seen in the 20% capacity plan.

## Data Availability

This article uses mathematical modeling and simulations, and does not use any real-world data generated from actual people.

